# Examining the Association between the COVID-19 Pandemic and Self-Harm Death Counts in Four Canadian Provinces

**DOI:** 10.1101/2021.10.13.21264961

**Authors:** Shelly Isnar, Mark Oremus

## Abstract

Governments implemented lockdowns and other physical distancing measures to stop the spread of SARS-CoV-2 (COVID-19). Resulting unemployment, income loss, poverty, and social isolation, coupled with daily reports of dire news about the COVID-19 pandemic, could serve as catalysts for increased self-harm deaths (SHD). This ecological study examined whether observed SHD counts were higher than predicted SHD counts during the pandemic period in the Canadian provinces of Alberta, British Columbia, Ontario, and Québec. The study also explored whether SHD counts during the pandemic were affected by lockdown severity (measured using the lockdown stringency index [LSI]) and COVID-19 case numbers. We utilized publicly available SHD data from January 2018 through November 2020, and employed AutoRegressive Integrated Moving Average (ARIMA) modelling, to predict SHD during the COVID-19 period (March 21 to November 28, 2020). We used Poisson and negative binomial regression to assess ecological associations between the LSI and COVID-19 case numbers, controlling for seasonality, and SHD counts during the COVID-19 period. On average, observed SHD counts were lower than predicted counts during this period (*p* < 0.05 [except Alberta]). Additionally, LSI and COVID-19 case numbers were not statistically significantly associated with SHD counts.

## 1. Introduction

SARS-CoV-2 is a highly contagious, zoonotic virus that was first discovered in Wuhan, China in late 2020 (Umakanthan et al., 2020). COVID-19 is the respiratory disease emerging from SARS-CoV-2 infection – symptoms include cough, fever, shortness of breath, anosmia, dysgeusia, chest pain, sputum production, and fatigue (Adil et al., 2021; Cabrera, 2021). According to the World Health Organization (WHO), approximately 13% of COVID-19 cases are severe, with an additional 6% of cases requiring hospitalization and intensive care (Adil et al., 2021). As of early October 2021, the cumulative number of confirmed deaths from COVID-19 globally is 608.52 per million people (Ritchie et al., 2021).

The spread of SARS-CoV-2 radically altered the everyday lives of people around the world, as they had to adjust to lockdowns and a range of other physical distancing measures, including the closure of non-essential businesses. In Canada, these closures resulted in an unemployment rate of almost 20% in January 2021, ten months after the start of the pandemic. This was the highest unemployment rate in over 10 years, with the exception of April 2020, when the unemployment rate was at 27% (Statistics Canada, 2021a). State-sponsored income replacement programs, when offered, barely exceeded the threshold of poverty for single persons in countries like Canada (MacEachen and Stahl, 2021).

Sudden loss of income and poverty are positively associated with anxiety, depression, and self-harm death (SHD) (Brenner and Bhugra, 2020; Funk et al., 2012; Kerr et al., 2017). These outcomes may be exacerbated by social isolation and loneliness arising out of lockdowns and physical distancing measures (Rothman and Sher, 2021), as well as by intense and often sensationalistic media coverage of new COVID-19 cases and intensive care unit (ICU) admissions (Georgiou, 2021).

This Canadian-based paper focused on SHD counts because SHD is one of the leading causes of death in Canada (Statistics Canada, 2021b). Therefore, clinicians and policymakers should understand the impact of COVID-19 and related risk factors on SHD, especially since COVID-19 is becoming endemic (Lavine et al., 2021). We defined SHD as: (1) any act or behaviour leading to a fatal outcome; (2) a self-initiated act or behaviour; and (3) an act or behaviour performed with the intention or expectation of bringing about death (Andriessen, 2011). Several papers reported increases in SHD following the start of the pandemic (Dsouza et al., 2020; Hill et al., 2021; Jenkins et al., 2021; Okubo et al., 2021; Sakamoto et al., 2021).

In contrast, several studies found that SHD during COVID-19 remained stable or decreased relative to pre-COVID-19 periods. Leske et al. (2021) used data from a real-time surveillance system and suicide register to compare Australian SHD rates in 2020 with those between 2015 and 2019. The overall rate of suspected SHD did not change after the declaration of the COVID-19 pandemic (14.07 deaths per 100,000 people from February 1, 2020 onward), compared to the pre-pandemic period (14.85 deaths per 100,000 people from January 1, 2015 to January 31, 2020).

Ueda et al. (2021) utilized publicly available data from the National Police Agency of Japan (January 2017 to October 2020) to compare SHD trends before and after the beginning of the COVID-19 pandemic. They reported that SHD dropped when Japan was first exposed to the virus in early 2020, compared to the average number of SHD in the corresponding months between 2017 and 2019. However, the decline reversed itself in July 2020, especially among females, with SHD between July and October 2020 exceeding the numbers observed in the corresponding months between 2017 and 2019. The authors believed SHD impacted females to a greater degree than males because women (especially those aged < 40 years) experienced the highest rates of job and income loss during the pandemic.

Pirkis et al. (2021) looked at SHD across 16 high-income and 5 upper-middle income countries during the earlier stages of the pandemic (April 1 to July 31, 2020). The authors reported that SHD risk did not increase during this period and remained stable in some countries, while rates in other countries decreased compared to expected rates. Other studies observed similarly stable or decreasing SHD rates during COVID-19 relative to pre-COVID-19 periods (Faust et al., 2021; Isumi et al., 2020, Karakasi et al., 2021; Kim, 2021; Leske et al., 2021; Li & Mitchell, 2021; Mehlum & Qin, 2021; Olie et al., 2021; Pirkis et al., 2021; Radeloff et al., 2021). These additional studies are summarized in Supplementary file 1.

### 1.1 Research Context

Only one study has investigated the impact of the COVID-19 pandemic on SHD in Canada (McIntyre et al., 2021). The authors reported a reduction in suicide mortality rate from 10.82 deaths per 100,000 between March 2019 and February 2020, to 7.34 deaths per 100,000 between March 2020 and February 2021. Given the importance of SHD in the Canadian context, coupled with mixed results in the literature and limited findings for Canada, additional work is required to understand trends in SHD on account of COVID-19. Additionally, to our knowledge, no study has examined risk factors for SHD during the pandemic.

We examined SHD counts in four Canadian provinces, i.e., Alberta, British Columbia, Ontario, and Québec, to assess whether SHD trends changed during a pandemic period from mid-March 2020 through November 2020, compared to a pre-pandemic period from January 2018 through mid-March 2020. We also examined whether lockdown severity and COVID-19 case numbers were associated with SHD counts between mid-March and November 2020.

## 2. Methods

### 2.1 Data Sources

Population-level SHD data for Alberta, British Columbia, Ontario, and Québec were assembled from the Canadian Vital Statistics Death database (Statistics Canada, 2021c). This database compiles all demographic and medical information from vital statistics registries across the Canadian provinces and territories.

The dataset containing COVID-19 cases in Ontario was assembled at the Public Health Unit (PHU) level and reported in the publicly accessible Ontario Data Catalogue (2021). We obtained COVID-19 case data for the other three provinces from data assembled by the Public Health Agency of Canada and reported by Statistics Canada (2021d). These data included COVID-19 case information such as region, episode week, age group, occupation, presence of symptoms, onset week of symptoms, hospitalization and ICU status, recovery time, death, and transmission.

Our measure of lockdown stringency came from the Oxford COVID-19 Government Response Tracker (Hale et al., 2021). Since January 2020, the tracker has amassed publicly available data from over 180 countries on items such as school and workplace closures, restrictions on public gatherings and travel, and vaccination policies. The lockdown stringency index (LSI) is a component of the tracker that quantifies the strictness of policies impeding citizens’ behaviour (e.g., restrictions on public gatherings). The LSI is expressed on a 0 to 100 scale, with higher values representing greater stringency.

### 2.2 Data Analysis

#### 2.2.1 Time-series

To display trends of provincial SHD counts over time, we plotted time (X-axis) versus SHD counts (Y-axis) on a weekly basis from January 6, 2018 to November 28, 2020, separately for each province. We transformed these data into an individual time-series object for each province, with one observation per week over the aforementioned period. The Dickey-Fuller test was used to assess the stationarity of the time-series data; we followed Prabhakaran’s (2021) procedure to transform the data to stationarity, if necessary.

#### 2.2.2 AutoRegressive Integrated Moving Average Modelling

We used AutoRegressive Integrated Moving Average (ARIMA) models and the observed SHD data from January 6, 2018 through March 14, 2020 to predict SHD during the COVID-19 period from March 21, 2020 through November 28, 2020. We initially examined whether the ARIMA model for each province would fit the complete set of observed SHD counts between 2018 and 2020. We employed the auto-ARIMA function to suggest the best-fitting ARIMA models for all predictive analyses.

For the period between March 21, 2020 and November 28, 2020, the weekly observed SHD counts were compared to the predicted counts generated by the ARIMA models. We employed Bland-Altman plots to show the average mean difference between observed and predicted SHD counts over the COVID-19 period.

#### 2.2.3 Regression

We employed regression analysis to explore the ecological association between (1) the LSI and COVID-19 cases as independent variables and (2) observed SHD counts between March 21, 2020 and November 28, 2020 as the dependent variable. We regressed SHD counts on the independent variables in a separate multivariable Poisson regression model for each province. For all models, we calculated robust standard errors, adjusted for seasonality, and tested for overdispersion using Cameron and Trivedi’s (1990) method (*p* < 0.05 suggested overdispersion). We ran negative binomial models in place of overdispersed Poisson models. Multicollinearity was assessed using variance inflation factors.

To conduct the analyses, we used R v4.0.4 (The R Project for Statistical Computing, Vienna, Austria) and the following R packages: AER, BlandAltmanLeh, forecast, lubridate, MASS, sandwich, and tseries. The level of statistical significance was set at α = 0.05. Ethics approval was not required for this study because the data were publicly available and reported at the aggregate (not individual) level.

## 3. Results

### 3.1 ARIMA

The predicted trends in SHD counts from the ARIMA models generally matched the observed SHD counts across the entire range of data from 2018 to 2020. The exception was for Alberta, where predicted trends increased slightly at the very end of the date range, while observed trends decreased (Figure 1). Using the pre-COVID-19 data (January 2018 through mid-March 2020) to predict SHD counts in the COVID-19 period, the ARIMA models predicted increasing trends for SHD counts in Ontario, decreasing trends in British Columbia and Alberta, and a roughly stable trend in Québec, compared to the pre-COVID-19 period (Figure 2).

**Figure 1:**
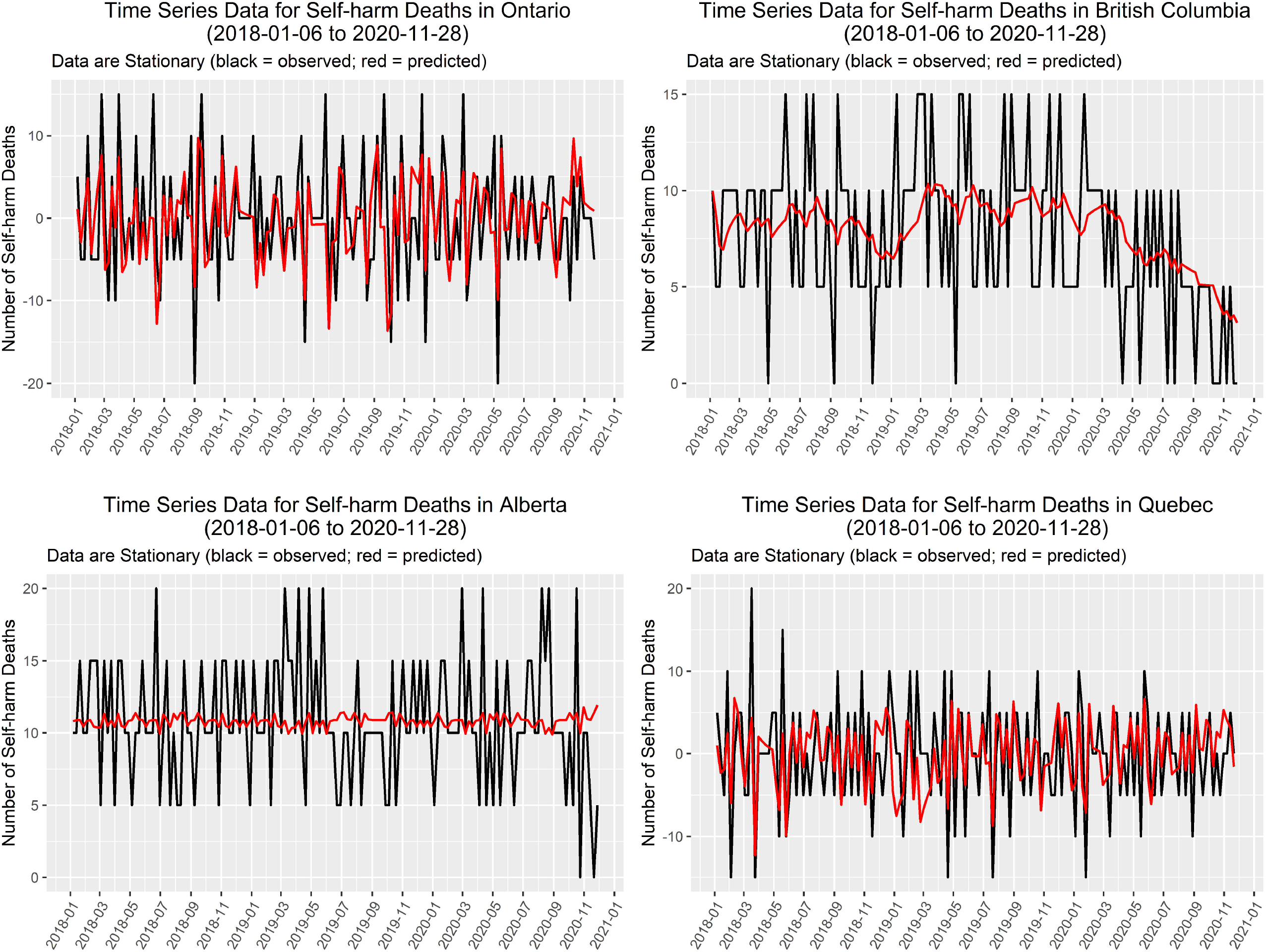
Observed versus Predicted Time Series Data for Self-harm Death Counts

**Figure 2:**
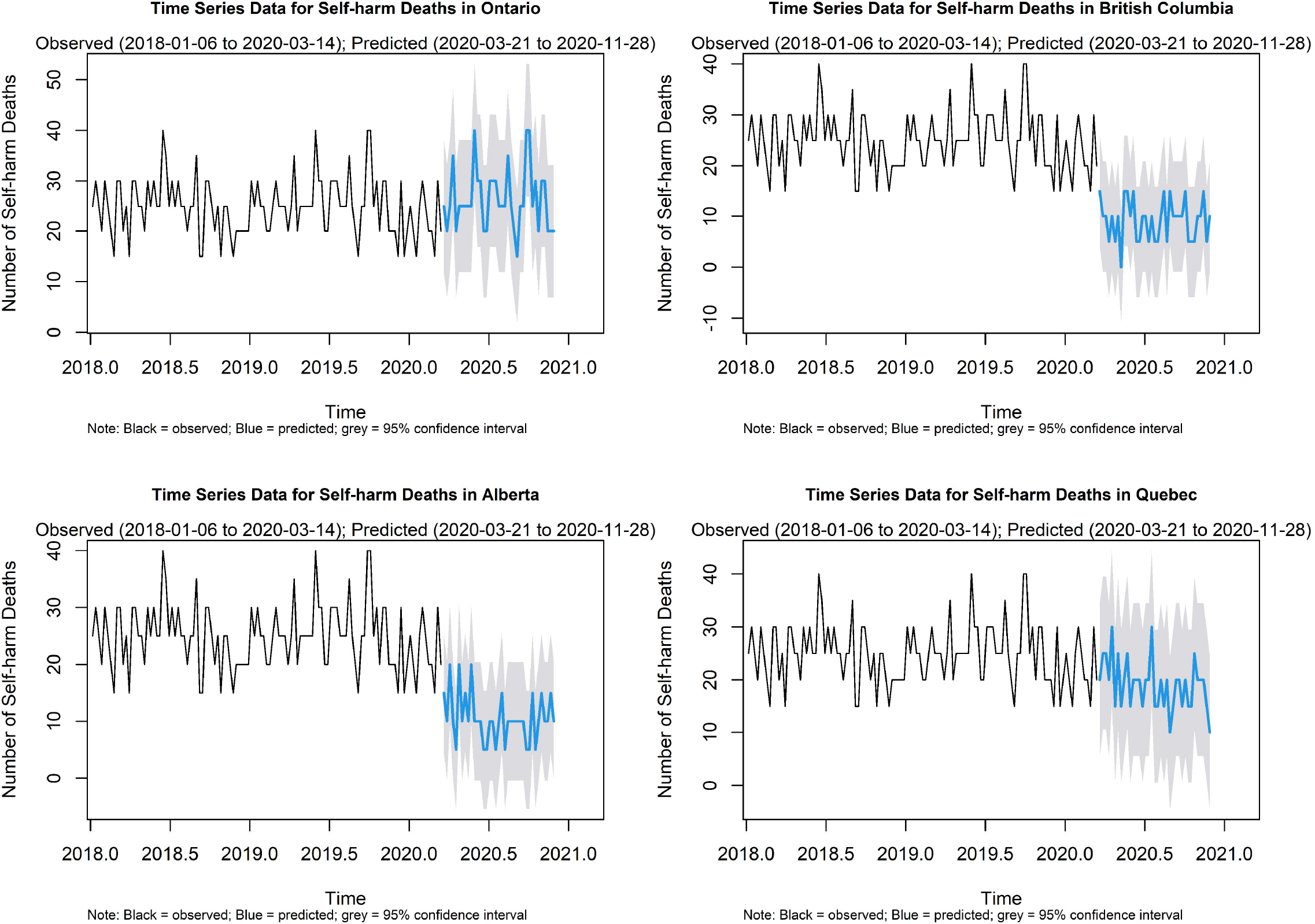
Predicted Self-harm Death Counts for COVID-19 Period based on pre-COVID-19 Self-harm Death Counts

### 3.2 Differences between observed and predicted self-harm deaths

On average, the actual, observed SHD counts during the COVID-19 period were lower than the predicted counts from the ARIMA models. The mean differences (SHD_observed_ – SHD_predicted_) were −10.8 (95% confidence interval [CI]: −13.52, −8.10), −4.73 (95% CI: −6.44, − 3.02), and −8.65 (95% CI: −10.82, −6.47) for Ontario, British Columbia, and Québec, respectively. The mean difference for Alberta was a minute −0.54 (95% CI: −2.99, 1.91). The Bland-Altman plots show that a majority of the weekly differences fell within two standard deviations of the mean differences (Figure 3).

**Figure 3:**
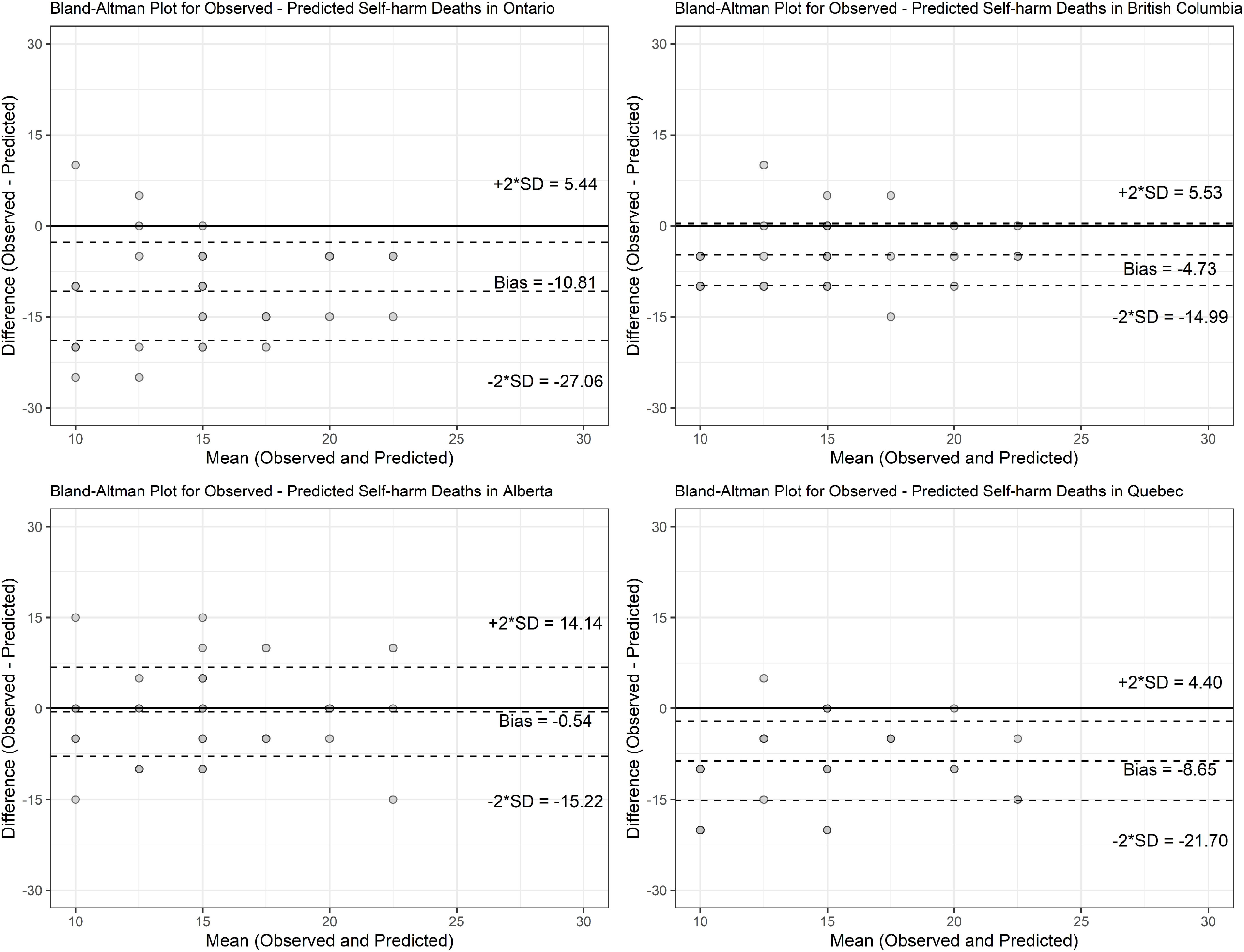
Bland-Altman Plots for Observed versus Predicted Self-harm Death Counts during the COVID-19 Period

### 3.3 Regression

The Poisson regression models for Alberta and British Columbia were overdispersed, so we utilized negative binomial models for these two provinces. Initially, we wished to include numbers of intensive-care unit (ICU) admissions in our regression models, but the variance inflation factors indicated that including both COVID-19 cases and ICU admissions in the same models led to multicollinearity. As such, we chose to remove ICU admissions and keep COVID-19 cases in the models because the average individual would be more likely to hear or read about case numbers than ICU admissions.

The regression models yielded similar findings across the four provinces (Table 1). In Ontario and Alberta, the baseline risks of SHD were approximately 16 and 15 cases per week, respectively, in the absence of COVID-19 cases, an LSI of 0, and the season being Fall. The baseline risks of SHD in Québec and British Columbia were about 9 and 4, respectively, under the same conditions.

**Table 1.**
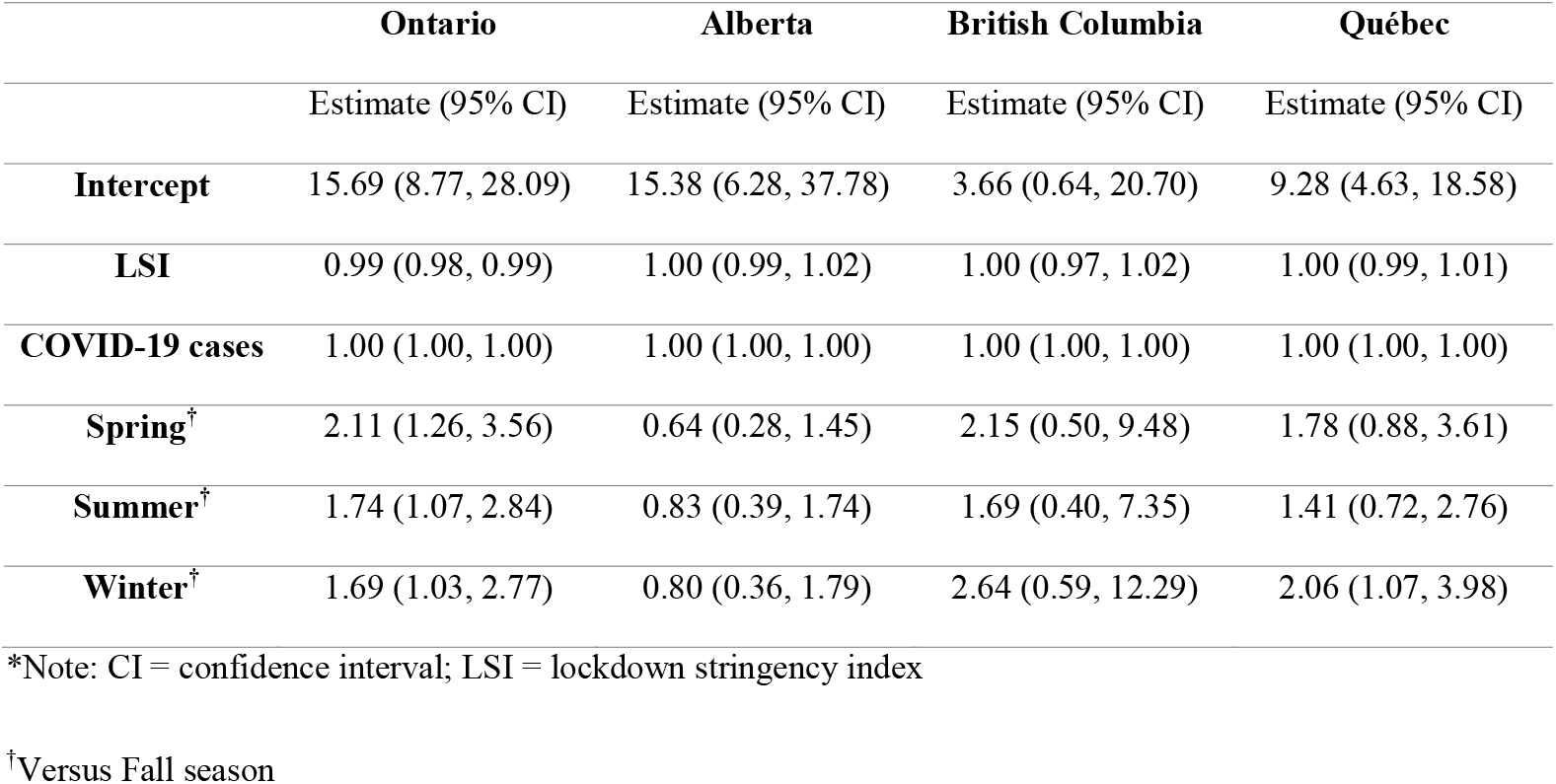
Multivariable Regression Results

In all four provinces, decreases in baseline SHD cases were negligible for each one-unit increase in COVID-19 cases when keeping all other variables constant; this was also the case for lockdown stringency, where the baseline SHD count decreased by approximately 1% for each one-unit increase in the LSI. Thus, a drastic increase in the LSI from 0 to 100 would only be expected to reduce baseline SHD by one person ([1 – 0.99] x 100).

## 4. Discussion

### 4.1 Observed versus Predicted Self-harm Deaths

Our findings were consistent across all four Canadian provinces. The observed mean SHD counts during the COVID-19 period were generally lower than the predicted mean SHD counts during the same period. A variety of factors may explain these findings. Isumi et al. (2020) suggested that lockdown measures provided an opportunity for family members to build stronger connections as a result of living in the same space for extended periods of time. This may have improved levels of functional social support for persons at risk of SHD, thereby reducing the incidence of SHD.

On occasion, the family dynamic may not be harmonious and family dysfunction could reduce social support and increase the risk of SHD. However, families who share space for extended periods might try harder to resolve familial conflicts and maintain peaceful living arrangements, thereby mitigating family dysfunction and the risk of SHD (Clark et al., 2016).

Ueda et al. (2021), Mehlum and Qin, (2020), and Radeloff et al. (2021) believed heightened social connectedness during the early stages of the pandemic led to lower rates of SHD in some localities. This connectedness may have reflected increased levels of altruistic behaviour that are often observed during times of natural disasters, with such behaviour being protective against SHD (Matsubayashi et al., 2013). Further, Faust et al. (2021), Pirkis et al. (2021), and Leske et al. (2021) thought the absence of increased SHD during COVID-19 may have resulted from newly implemented community supports for persons struggling with mental health during the pandemic.

### 4.2 Regression Analysis

Across the four provinces, LSI and COVID-19 case numbers had little or no effect on SHD. Perhaps improved familial relationships, social connectedness, or other diversions (e.g., binge-watching internet streaming services [Boursier et al., 2021]) counteracted the adverse impacts of lockdown measures or public attention given to COVID-19 case counts. Interestingly, we did not find any published studies looking at associations between LSI/COVID-19 case counts and SHD.

### 4.3 Strengths and Limitations

The major strength of this study is the utilization of publicly available, population-level data from four Canadian provinces to conduct the analyses. These data spanned the pre-COVID-19 and COVID-19 periods, thereby allowing us to compare SHD counts before and after the pandemic’s effects were felt in Canada. However, since ex post facto interviews are impossible to conduct on persons who suffer a SHD, we could not examine the extent to which lockdowns or COVID-19 case numbers may have influenced specific SHDs. Further, SHD data for 2020 could underreport true SHD counts as a result of possibly misclassifying SHD as accidental poisonings, alcohol-induced deaths, or illicit drug deaths (McIntyre et al., 2021).

### 4.4 Policy Suggestions

While we did not find evidence of a COVID-19 impact on SHD, accessibility to mental health services should continue to be a health priority. The various factors responsible for reducing SHD during the pandemic may become less important as COVID-19 becomes endemic and societal structures re-open. Thus, SHD may eventually climb to pre-pandemic levels and mental health services will be required to address the increase.

## 5. Conclusion

In this study, we analyzed population-level data from four Canadian provinces and found observed SHD counts were lower than predicted counts during COVID-19. Also, neither LSI nor the number of COVID-19 cases was statistically significantly associated with SHD. Future research should continue to explore the reasons for reduced SHD counts during COVID-19 and identify a greater range of possible risk and protective factors for SHD in the COVID-19 era.

## Data Availability

All data produced are available online from the Ontario Data Catalogue and Statistics Canada.

## Acknowledgements

We thank Jackie Stapleton for devising the literature search used to retrieve background articles for this paper.

## Supplementary File

Supplementary File 1 – Summary of Studies on Self-harm Deaths Before and After the COVID-19 Pandemic

