## Supplemental File 1 for "Examining the Association between the COVID-19 Pandemic and Self-Harm Death Counts in Four Canadian Provinces"

**Supplementary File 1 – Summary of Studies on Self-harm Deaths Before and After the COVID-19 Pandemic**

According to Kim (2021), self-harm death (SHD) rates in Korea decreased by 6.9% in the first eight months of 2020 compared to the same period in 2019. Male SHD decreased by 10.1%, while female SHD showed a 1.4% increase. After March 2020, when the number of COVID-19 deaths and cases were still at their peak, males showed significantly decreased SHD, while females showed increases in March and April 2020, and a decrease in May 2020.

Li and Mitchell (2021) examined coroner’s files to assess SHD rates among the elderly and racial minorities during strict, state-wide lockdowns in Connecticut from March 10 to May 20, 2020. Data from this period were compared to the matching date range during the previous six years of SHD data. Seventy-four completed SHD were reported in Connecticut during the lockdown period (age-adjusted rate of 9.4 per 100,000 person-years). This rate was 20% lower than the previous year and 13% lower than the recent 5-year average of 10.8 per 100,000 person-years.

Radeloff et al. (2021) used the City of Leipzig’s cause of death statistics from 2010 to 2020 to assess SHD during periods with and without restrictions on freedom of movement and social interaction during the pandemic. Results showed mean SHD per 100,000 life-years were lower in periods with severe COVID-19 restrictions (-7.2) compared to periods without restrictions (i.e., 7.2 versus 16.8 [*p* = 0.045]) in 2020. When compared to previous years, the authors found that difference was due to unusually high SHD prior to implementing restrictions, while SHD during the pandemic were within the ‘normal’ range for previous years (expected suicides = 32.3, observed suicides = 35; incidence rate ratio [IRR] = 1.084, *p* = 0.682).

Calderon-Anyosa and Kaufman (2021) used data from the Peruvian National Death Information system to conduct an interrupted time series analysis stratified by sex and a time unit of every 15 days from January 1 2017 to September 26 2020. The lockdown period was defined as starting from March 16 2020 for which trends in SHD were assessed in comparison to the same time period in 2017-2019. Results suggested a 1 per million decrease in female SHD and a 2 per million decrease in male SHD during the pre-lockdown period. Post-lockdown SHD for men showed an upward trend in slope compared to SHD in previous years (an additional 1.20 deaths per million men per year [95% confidence interval (CI): −0.26, 2.65]).

A population-based time series conducted by Faust et al. (2021) looked at SHD data from Massachusetts vital records between January 2015 and May 2020. The incidence rate of suicide deaths in Massachusetts was 0.67 (95% CI, 0.56, 0.79) per 100,000 person-months during the pandemic, compared to 0.80 (95% CI, 0.68, 0.93) per 100,000 person-months during the same period of the previous year, in persons aged 10 years or over. Furthermore, results also suggested that the demographics and characteristics of suicide deaths during the pandemic did not deviate from corresponding times of the year between 2015 and 2019 (Faust et al., 2021).

Isumi et al. (2020) assessed monthly SHD during the lockdown (March to May 2020) among persons under 20 years old in Japan using public suicide statistics to assess any changes for the same time period in 2018-2019. SHD rates were not found to significantly change during periods of school closure during the pandemic. However, it was found that suicides increased significantly in May 2020 compared to March 2020 (IRR = 1.15; 95% CI: 0.81, 1.64).

A study conducted in Norway by Mehlum and Qin (2021) examined death registry data from March to May 2020 and found that SHD during this timeframe were lower than when compared to the previous 5 years. There were 140 SHD counted from March to May 2020 (2.8 per 100,000) compared to an average of 165 SHD per year from 2014 to 2018.

Lastly, a study conducted in Greece by Karakasi et al. (2021) also showed no increase in SHD during 2020 when compared to previous years. The researchers compared the period from March 1 to May 15, 2020, to the same date periods between 2010 and 2019 and found no significant differences in the method of death across the study periods (Fisher exact test, *p* > 0.05).

*Note:* References are shown in the ‘References’ section of the paper.
